# The effect of focused muscle contraction therapy on chronic pain and Brodmann Area activity in former National Football League players

**DOI:** 10.1101/2022.03.09.22272106

**Authors:** Neli Cohen, Greg Hachaj, Jose Rubio, Alexandra Kastelz, Marcin Hachaj, Dan Zierfuss, Maab Osman, Pete Tsiampas, Bo Fernhall, Effrossyni Votta Velis, Enrico Benedetti, Amelia Bartholomew

**Affiliations:** Department of Surgery, University of Illinois; GhFitlab, Chicago; College of Applied Health Sciences, University of Illinois; Holistic Neuropsychology, Chicago

## Abstract

NFL players have a traumatic injury rate approaching 100%; chronic pain with decreased concentration occur commonly. This study examined the role of a novel focused muscle contraction therapy for the treatment of chronic pain and identified its impact on brain activity. Chronic pain was assessed by numerical score, neuropathic component, and impact on daily activities in 8 retired players. Brain activity was characterized by QEEG with low-resolution electromagnetic tomography analysis and functional measures of visual and auditory attention. Focused muscle contraction muscle therapy administered twice weekly for 6 months was tapered to twice monthly by 12 months. Brodmann Areas (BA) 4 and 9, known to associate with chronic pain, showed values outside the clinically normal range; mean pain duration was 16.5 ± 12.9 years. At 6 months, 5/8 subjects reported pain scores of 0. High beta wave activity was seen in BA 19, 21, 29, 30, and 39, affecting auditory, visual, and body perceptions. Clinically relevant improvements were observed in auditory attention and visual stamina. Pain relief was sustained through 18 months of follow-up. Focused muscle contraction therapy appears to redirect brain activity to new areas of activity which are associated with long-lasting relief of chronic pain and its detriments. This study was registered with clinicaltrial.gov #NCT04822311.

## INTRODUCTION

For National Football League (NFL) players, the injury rate is nearly 100% and is associated with long-term physical and cognitive disabilities. Patients who suffer from injury-related chronic pain can experience pain in multiple locations for several years after injury leading to difficulties with sleeping, fatigue, depression, and low quality of life, the lowest of any medical condition [6; 11; 26; 32; 41]. Treatment strategies for chronic pain include anti-depressants, anti-convulsant, topical agents, injection therapies, physical therapy, and psychological approaches, but none provide more than a 30% reduction in pain [17; 23; 50].

Exercise has been recommended for and used in the treatment of chronic pain [17; 45]. This recommendation can be at odds with the patient in pain who may be fearful to undertake any movement that may aggravate their pain; fear of pain and subsequent reduced activity/disuse plays a role in the transition from acute to chronic pain [47; 51]. Ideally, the choice of physical activity must be tolerable, encourage continued engagement, and evoke positive feelings.

Chronic pain changes the structure of the brain leading to decreased cortical gray matter in the bilateral dorsolateral prefrontal cortex (Brodmann areas 46 and 9), thalamus, brainstem, the primary somatosensory cortex (Brodmann areas 1,2,3), and posterior parietal cortex (Brodmann area 5) [2; 8; 30; 44]. The duration of pain and its intensity can predict regional atrophy in the dorsolateral prefrontal cortex [2]. Eliminating chronic pain is associated with restoration of brain anatomy and function [24].

The areas of the brain which are diminished in form and function in the presence of chronic pain are also involved in the successful learning of new motor tasks [4]. The dorsolateral prefrontal cortex is also highly engaged with learning new motor tasks. Engagement decreases when a motor task becomes more “automatic”; it is believed that transition to automatic function is delegated to lower brain structures [13; 20]. Most exercise regimens become routine and automated; automatic activities [43; 55; 56] have not been effective in treating chronic pain or restoring anatomic form and function [52]. These lower brain structures are not involved in chronic pain. Eliciting a deliberate contraction of a specific muscle requires intensive concentration and is most effective if accompanied by verbal instruction [46]. Such concentration implies the need for higher-order brain function, such as the dorsolateral prefrontal cortex. Active stimulation in a focused motor task may stimulate areas of the brain affected by chronic pain, restoring brain anatomy and function and, as a consequence, extinguish the perception of chronic pain.

In our previous randomized clinical trial of focused muscle contraction therapy, 80 seriously ill kidney transplant recipients demonstrated improved measures of global physical health (P = 0.0034), global mental health (P=0.0064), and elimination of chronic pain [22]. The purpose of this study was to examine the role of focused muscle contraction as a treatment for chronic pain and to identify the role of focused muscle contraction on brain activity associated with chronic pain.

## METHODS

### Study Design

The study was submitted to ClinicalTrials.gov and was registered as trial # NCT04822311. In consideration of clinical trial design recommendations set forth by IMMPACT, (Initiative on Methods, Measurement, and Pain Assessment in Clinical Trials) this study was conducted as a longitudinal pilot in which duration of treatment was 18 months; 6 months of twice-weekly 60-minute sessions, followed by 6 months of weekly 60-minute sessions, and maintenance therapy thereafter with twice-monthly 60-minute sessions.

### Study participants

*Sample Size, Inclusion, and Exclusion Criteria*: A sample size calculation of 20 was expected to reduce the analog pain score 3 increments, assuming a two-tailed test using an alpha of 0.05 and a power of 0.8. Eight retired football players were enrolled with such dramatic results, that it was decided to provide a report on the findings before completion of enrollment. Focused muscle contraction therapy was initiated with ultra-low weights (see **Supplementary Table 1**). Participants were excluded if they had any presence of tumor, acute fracture, history of surgery in the past 2 months, inability to travel to undertake therapy, or inability to give consent. Subjects were eligible if they 1) played professional football in the NFL and 2) suffered from chronic pain lasting a minimum of 6 months with a minimum severity of 5 out of 10 points on the Numeric Pain Rating Scale and 3) had failed standard treatment. All subjects signed informed consent and the study was approved by the UIC IRB.

### Focused Muscle Contraction

The intervention incorporated a specially designed, low intensity, resistance-based exercise regime (GH Method) as previously reported [49]. Exercise prescribed were biceps curl, chest press, shoulder press, triceps extension/pushdown, lat pull-downs, front row, sit-ups, leg extension, leg curl, leg press, leg abduction, leg adduction. In the first 4-8 sessions, the intensity was low to avoid fatigue and pain. Therapy was initiated with one 30 second set of exercises using 2-second concentric and 3-second eccentric contraction for each exercise. Verbal instruction was coupled with tactile identification of the muscle group to ensure focused contraction was being accomplished for the specific muscle/muscle group. Verbal encouragement was given with correctly performed contractions while tactile reminders, administered by gently tapping on the muscle to be contracted, were given if the contraction was not adequately executed. Exercise intensity was prescribed at a Fatigue score of 3 or below (indicating mild fatigue) using the 10-point Fatigue Scale. In the second phase, the emphasis was on progressively developing muscle strength and function of the major muscle groups. The first included 15–20 repetitions at a perceived exertion of 3 or less using the 10-point Borg Scale of Perceived Exertion. The Borg scale has been validated for use during resistance exercise training [42; 49]. For twice-weekly therapy, day one included 10–12 repetitions at a perceived exertion of 3–4 on the Borg Scale and 6–8 repetitions on day 2 of each week, at a perceived exertion of 4–5. Patient fatigue and pain levels were monitored throughout the exercise session using the 10-point fatigue scale, without reaching above a 4 rating. This phase lasted 8–10 weeks depending on the patient progression and capability. In the third phase, the emphasis was on the further development of muscle strength, function, and endurance utilizing both major and smaller muscle groups. The intensity was progressively increased based on the individual response of each patient. All exercises were done without going above 4 on the Fatigue Scale. Phase 3 continued for the remainder of the study. Before each session, a 5-to 10-minute light warmup plus stretching was incorporated and pain scores were obtained. These sessions were supervised by trained personnel.

### Pain intensity

The Numeric Rating Scale for Pain, in which the subject selected the number from 1-10 with 10 being the worst pain experienced, was used to measure the intensity of pain before treatment, and then before every treatment session thereafter. The DETECT questionnaire for the detection of neuropathic pain was measured pre-treatment and at 6 months.

### Impact of pain

The impact of pain was characterized by the number of years of persistence, the number, and type of failed therapies, and the effect on sleep, eating habits, and mood.

### Measures of Cognitive Function

Cognition was measured using the Integrated Visual and Auditory Scale (IVA-2). This measures visual and auditory attention and response control functioning [14]. Outcomes measured included vigilance, focus, speed, prudence, consistency, stamina, comprehension, persistence, and sensory/motor responses.

### QEEG

The Quantitative Electroencephalogram (QEEG) provides a highly significant predictive relationship of self-reported pain scores [40]. The QEEG was used to localize and quantify the functional systems the brain was using compared to age-matched normative ranges before and 6 months after intervention. The content validity of the qEEG has been established by correlation with MRI, PET, SPECT, the Glasgow Coma Score, and others [48]. A dry electrode cap with premeasured sites with the international 10/20 system of electrode placement was fitted to each subject and consisted of 19 electrodes placed in the following positions: FP1. F3, C3, P3, O1, F7, T3, T5, FP2, F4, C4, P4, O2, F8, T4, T6, Fz, Cz and Pz. The EEG was digitally recorded using the WinEEG (Mitsar, St Petersburg, Russia) acquisition software utilizing 19 electrodes referenced to A1 and A2 (linked ear montage). The input signals were amplified (bandpass 0.3– 70 Hz) and sampled at the rate of 250 Hz. The EEG was recorded continuously in the awake state with eyes closed and, in the waking, relaxed state eyes-open condition; the participants were sitting in an upright relaxed position and given instructions to remain still, inhibit eye movements, blinks, and muscle activity from the forehead neck and jaws. A trained neuropsychology technician monitored each subject during the recording and ensured the subject was awake. EEG data were collected in accordance with accepted standards of QEEG acquisition methods to ensure quality recordings. The absolute power, relative power, amplitude asymmetry, coherence, and phase lag for surface potentials were digitally filtered and analyzed for eight frequency bands: delta (1–4 Hz), theta (4–8 Hz), alpha (8–12 Hz), beta1 (12– 15 Hz), beta2 (15–18 Hz), beta3 (18–25 Hz), and high beta (25–30 Hz). Using QEEG, the functional systems that the brain is using compared to age-matched normative ranges before and after the intervention was localized and quantified.

### LORETA

QEEG findings were used to identify active areas of the brain using low-resolution electromagnetic tomography (LORETA) [37; 38]. LORETA is an inverse solution technique that estimates the distribution of electrical neuronal activity in a 3-dimensional space. It has been extensively used in electrophysiological research [36] and evaluated independently in several laboratories [16; 53; 54]. LORETA is a useful tool to investigate the location of specific oscillation frequencies of brain activity known as Brodmann Areas. Previous studies that compared LORETA with fMRI results, showed that LORETA was equally capable of localizing single and multiple active brain activities and was in congruence with fMRI measures [34].

### Data collection and management

All study personnel underwent training on data safety monitoring and subject confidentiality issues. Each subject was assigned a Study Identification Number and the data were collected by study number and then entered into a password-protected database for analysis. All data was collected pre-treatment and at 6 months with numerical pain scores collected before every therapy session through 18 months. The research team was responsible for all data analyses.

### Statistical Analysis

The statistical method for determining significant changes in QEEG measures following treatment was based on the method developed by Wigton and Krigbaum, 2015 [25]. Z-score values were provided by the QEEG software for each of the 19 channels per brain wave per subject by Absolute Power and by Relative Power. Each of the 64 paired channels per brain wave per subject was used to report asymmetry and coherence. An absolute value transformed z-score threshold of 1.5 was considered a clinically relevant site of interest (SOI) [19], which means that baseline z-scores for each of these channels were defined as SOIs if they were farther than +1.5 or -1.5 standard deviations from the normative mean. For each channel, the SOIs were averaged for each individual pre-treatment and compared to the average post-treatment z-score values for corresponding SOIs that were used in the pre-treatment calculation. In this analysis, each individual had up to 19 average z-score values for pre-treatment SOIs in measurements of absolute and relative powers and had up to 64 average z-score values for pre-treatment SOIs for measures of coherence and asymmetry. Paired t-tests performed between the pre- and post-treatments average z-scores were used to evaluate mean changes in SOIs for each parameter and were considered statistically significant if the p-value<0.05. Changes in Z-scores which exceeded 1.5 from the mean as an SOI but then became less than 1.5 from the mean post-treatment indicated normalization and were no longer considered clinically significant.

## RESULTS

### Characteristics of Chronic Pain

Of the 8 retired male NFL football players enrolled, the mean age was 59.0 ± 13.8 years old and the mean a number of years in pain was 16.5 ± 12.9 years. **Table 1** presents initial characteristics reported at baseline. Participants were male, 62.5% were Black or African American and 37.5% were white. Half were married or in a domestic partnership. The pain was unrelenting and persisted for a minimum of five years with 75% experiencing chronic pain for 5-20 years. All participants reported failure of standard therapies with all subjects undergoing treatment with an average number of 7.8 ± 6.1 treatments. Some subjects (n=4) undertook as many as 8 different therapies. All subjects reported at least one type of sleep disturbance, 72.5% reported an effect on appetite, and 62.5% reported an effect on mood.

**Table 1.** Demographics.

| Categories | Incidence |
| --- | --- |
| **Ethnicity** |  |
| Black or African American | 62.5% |
| White | 37.5% |
| **Marital status** |  |
| Married or domestic partnership | 50% |
| Single, never married | 25% |
| Divorced | 12.5% |
| Widowed | 12.5% |
| **Length of pain** |  |
| 5-10 years | 37.5% |
| 11-20 years | 37.5% |
| 21-45 years | 25% |
| **Surgery** |  |
| Yes | 75% |
| No | 25% |
| **Pain effect on appetite** |  |
| Decreased appetite | 25% |
| Increased appetite | 12.5% |
| Not effect on appetite | 37.5% |
| Gained weight | 25% |
| **Pain effect on sleep** |  |
| Sometimes disturbs sleep | 50% |
| Trouble falling asleep/Pain wakes me up | 50% |
| Less than 7 hours sleep due to pain | 50% |
| **Pain effect on mood** |  |
| Stressed | 50% |
| Nervous | 25% |
| Less interest in activities | 62.5% |
| Sad or blue | 12.5% |
| **Drugs or treatments** |  |
| NSAIDs | 62.5% |
| TENS | 25% |
| Acupuncture | 37.5% |
| Chiropractic Therapy | 75% |
| Occupational Therapy | 25% |
| Ultrasound Therapy | 50% |
| Massage Therapy | 87.5% |
| Physical Therapy | 75% |

The neuropathic component of pain was measured by the DETECT questionnaire, **Table 2**. Subjects graded their pain on a scale from 0 (hardly noticed) to 5 (very strong pain). Every subject reported a component of neuropathic pain. The two most common complaints were a tingling sensation with a mean grade of 1.75 ± 1.4 and the sensation of numbness with a mean grade of 1.6 ± 1.4; both had an incidence of 6/8 subjects. The next most frequent characteristic was a burning sensation with an average score of 2.1 ± 2.1. The least frequent pain characteristic was the ability of light touch to elicit pain, and this was reported by only one subject. At 6 months, the same pattern of resolution of pain reflected in the numerical pain scores by site (**Table 3)**, was also reflected in the neuropathic pain for every subject with 5/8 subjects indicating the absence of chronic pain.

**Table 2.** Neuropathic Characteristics of pain *(0: never; 1: Hardly noticed; 2: slightly; 3: moderately; 4: strongly; 5: very strongly)*

| Gradation of pain | S1 | S2 | S3 | S4 | S5 | S6 | S7 | S8 |
| --- | --- | --- | --- | --- | --- | --- | --- | --- |
| Burning sensation | 4 | 4 | 1 | 0 | 0 | 0 | 3 | 5 |
| Tingling or prickling sensation | 1 | 4 | 2 | 2 | 0 | 0 | 3 | 2 |
| Painful light touching | 0 | 0 | 0 | 0 | 0 | 0 | 2 | 0 |
| Sudden pain attacks | 0 | 0 | 1 | 0 | 0 | 3 | 2 | 5 |
| Painful cold or heat | 0 | 0 | 0 | 0 | 2 | 0 | 2 | 2 |
| Sensation of numbness | 0 | 0 | 2 | 2 | 1 | 4 | 1 | 3 |
| Slight pressure | 0 | 0 | 2 | 0 | 4 | 2 | 0 | 1 |

**Table 3.** Pre and 6-months Post-therapy Numerical Pain Scores by Site and Subject Number. *(0: No pain; 10: Worst pain imaginable; *Difference between pre and post was statistically significant defined as p<*.*05)*

| Body part | Subject 1 |  | Subject 2 |  | Subject 3 |  | Subject 4 |  | Subject 5 |  | Subject 6 |  | Subject 7 |  | Subject 8 |  |
| --- | --- | --- | --- | --- | --- | --- | --- | --- | --- | --- | --- | --- | --- | --- | --- | --- |
|  | PRE | POST | PRE | POST | PRE | POST | PRE | POST | PRE | POST | PRE | POST | PRE | POST | PRE | POST |
| Neck | 9 | 0 | 5 | 0 |  |  | 7 | 0 | 5 | 2 | 5 | 0 |  |  | 4 | 0 |
| Trapezius |  |  |  |  |  |  |  |  |  |  |  |  | 5 | 0 |  |  |
| R.Shoulder | 7 | 0 |  |  | 8 | 0 |  |  |  |  |  |  |  |  | 2 | 0 |
| L.Shoulder |  |  |  |  |  |  |  |  |  |  |  |  | 8 | 0 | 7 | 0 |
| R. Elbow |  |  |  |  | 3 | 0 |  |  |  |  | 3 | 0 |  |  |  |  |
| L. Elbow |  |  |  |  | 3 | 0 |  |  |  |  |  |  |  |  |  |  |
| R. Hand |  |  |  |  |  |  |  |  |  |  |  |  |  |  |  |  |
| L. Hand |  |  |  |  |  |  |  |  |  |  | 8 | 1 |  |  |  |  |
| R. Wrist |  |  |  |  |  |  |  |  |  |  |  |  | 8 | 0 | 4 | 0 |
| L. Wrist | 7 | 0 |  |  |  |  |  |  |  |  |  |  | 8 | 0 |  |  |
| Upper Back |  |  | 4 | 0 |  |  |  |  |  |  |  |  |  |  |  |  |
| Lower Back |  |  | 7 | 0 |  |  | 9 | 0 | 4 | 0 | 7 | 0 | 6 | 0 | 7 | 0 |
| R. Knee | 8 | 0 | 5 | 2 |  |  |  |  | 6 | 0 | 5 | 0 | 8 | 0 | 6 | 0 |
| L. Knee |  |  | 5 | 0 | 6 | 0 |  |  | 8 | 0 |  |  |  |  |  |  |
| R. Hip | 6 | 0 |  |  |  |  | 3 | 0 |  |  |  |  |  |  |  |  |
| L. Hip |  |  | 6 | 1 | 8 | 3 |  |  |  |  | 4 | 0 |  |  |  |  |
| R. Foot |  |  | 5 | 0 |  |  |  |  |  |  |  |  |  |  | 7 | 0 |
| L. Foot |  |  | 5 | 0 |  |  |  |  |  |  |  |  |  |  |  |  |
| R. Toes |  |  |  |  |  |  |  |  |  |  |  |  |  |  |  |  |
| L. Toes |  |  |  |  |  |  |  |  | 3 | 0 |  |  |  |  |  |  |
| Calves (both) |  |  |  |  |  |  |  |  |  |  | 10 | 0 |  |  |  |  |
| Hamstrings |  |  |  |  |  |  |  |  |  |  |  |  |  |  | 5 | 0 |
| MEAN<br>(STD) | 7.4<br>(1.1) | 0.0<br>(0.0) | 5.3<br>(0.9) | 0.4<br>(0.7) | 5.6<br>(2.5) | 0.6<br>(1.3) | 6.3<br>(3.1) | 0.0<br>(0.0) | 5.2<br>(1.9) | 0.4<br>(0.9) | 6.0<br>(2.4) | 0.1<br>(0.4) | 7.2<br>(1.3) | 0.0<br>(0.0) | 5.3<br>(1.8) | 0.0<br>(0.0) |
| T-test<br>P-value | <.001* |  | <.001* |  | .004* |  | .022* |  | <.001* |  | <.001* |  | <.001* |  | <.001* |  |

Every subject demonstrated pain in more than one location, with an average of 5 +/-2 locations affected in each player, **Table 3**. The pain was reported in the following locations: head, neck, shoulders, wrists, back, knees, ankles, and feet. Before treatment, the mean pain score of the subjects was 6.0 ± 0.9 on a scale of 10, with 0 indicating the absence of pain and 10 indicating the worst pain imaginable.

After six months of muscle-contraction therapy, 5/8 subjects reported pain scores of 0 in all sites previously identified as painful. Subject 2, who reported substantive chronic pain in 8 different locations had a resolution of 6 sites to 0 with significant pain reduction from 5 to 2 of the right knees and 6 to 1 of the left hip. Subject 3 achieved scores of 0 in 4 of 5 locations with a reduction from 8 to 3 in chronic left hip pain. Subject 5 achieved a score of 0 in 3 of 4 sites with a reduction from 5 to 2 in chronic neck pain. The aggregate 6 months mean pain score was 0.2 ± 0.6 (p<.001 vs pre-treatment). The difference in pain relief between was also statistically significant for the average pain score for every subject individually. When all the pain scores the subject had in different body parts was averaged pre-therapy and compared to the average score in all body parts at 6 months post-treatment, all were significantly reduced or abolished, with p-values<0.05, **Table 3**. Pain cessation by the numerical score in most subjects was observed by 3 months of treatment. This effect remained stable through 18 months.

### QEEG parameters

Brain oscillation activity was reported by frequency as assigned by -delta, theta, alpha, and beta frequencies for each individual with eyes closed and with eyes open. Mean SOIs pre-treatment were compared to values measured 6 months post-treatment using paired t-tests, **Table 4**. The metrics for all frequency bands included absolute power (energy in a chosen frequency band), relative power (energy in a chosen frequency band divided by the total energy from all frequency band), coherence (a mathematical method that can be used to determine if two or more brain regions have similar neuronal oscillatory activity with each other) [5], and asymmetry [10]. The number of subjects who had at least one SOI for the given parameter for the given metric was represented by n in **Table 4**, with P indicating the p-value from the 2-sided paired t-test on differences between pre and post.

**Table 4.** Mean changes in positive and negative z-scores QEEG parameters for averaged sites of interest (SOIs) pre-treatment to post-treatment.

|  |  | EYES CLOSED |  |  |  |  |  |  |  | EYES OPEN |  |  |  |  |  |  |  |
| --- | --- | --- | --- | --- | --- | --- | --- | --- | --- | --- | --- | --- | --- | --- | --- | --- | --- |
|  |  | Mean Positive z-scores |  |  |  | Mean Negative z-scores |  |  |  | Mean Positive z-scores |  |  |  | Mean Negative z-scores |  |  |  |
| Metric | Parameter | n | Pre (SD) | Post (SD) | P | n | Pre (SD) | Post(SD) | P | n | Pre (SD) | Post (SD) | P | n | Pre (SD) | Post (SD) | P |
| Absolute Power | Delta | 3 | 2.0 (0.3) | 0.9 (1.1) | .003* | 2 | -1.9 (0.3) | -1.3 (0.9) | .021* | 3 | 3.5 (1.8) | 0.1 (1.1) | <.001* | 3 | -2.2 (0.4) | -1.3 (1.0) | <.001* |
|  | Theta | 2 | 1.8 (0.3) | 0.9 (0.6) | <.001* | 1 | -1.6 (0.1) | -1.6 (0.7) | .903 | 3 | 2.3 (0.9) | 0.5 (0.8) | <.001* | 3 | -2.2 (0.5) | -1.1 (1.0) | <.001* |
|  | Alpha | 2 | 1.7 (0.2) | 1.4 (0.5) | .368 | 2 | -1.5 (0.0) | 0.4 (1.1) | .011* | 2 | 1.8 (0.3) | 0.6 (0.9) | <.001* | 2 | -1.9 (0.2) | -0.6 (0.6) | <.001* |
|  | Beta | - | - | - | - | 1 | -1.7 (0.3) | -1.0 (0.4) | .079 | 2 | 2.8 (1.5) | -0.1 (1.6) | .201 | 2 | -2.5 (0.3) | -0.8 (0.6) | <.001* |
|  | High Beta | 1 | 1.7 (0.1) | 3.5 (1.2) | .162 | - | - | - | - | 3 | 2.2 (0.6) | 0.9 (0.6) | <.001* | 2 | -1.7 (0.1) | -1.1 (0.2) | <.001* |
|  | Beta 1 | - | - | - | - | - | - | - | - | 1 | 2.3 (1.4) | 0.7 (0.1) | .120 | 2 | -1.9 (0.2) | -1.0 (0.2) | <.001* |
|  | Beta 2 | - | - | - | - | - | - | - | - | 1 | 2.4 (1.1) | 0.4 (0.5) | .041* | 2 | -1.9 (0.2) | -1.1 (0.2) | <.001* |
|  | Beta 3 | - | - | - | - | - | - | - | - | 2 | 1.9 (0.6) | 0.9 (0.6) | .005* | 3 | -2.1 (0.3) | -1.1 (0.2) | <.001* |
| Relative Power | Delta | 2 | 1.7 (0.2) | 0.6 (1.0) | .079 | 1 | -1.7 (0.1) | -0.2 (0.6) | <.001* | 2 | 1.7 (0.3) | -0.2 (1.0) | <.001* | 3 | -1.9 (0.4) | 0.7 (1.2) | <.001* |
|  | Theta | 3 | 1.6 (0.1) | 0.5 (0.7) | <.001* | - | - | - | - | 2 | 1.9 (0.2) | 0.3 (0.8) | <.001* | 1 | -1.7 (0.2) | -0.6 (0.7) | .173 |
|  | Alpha | - | - | - | - | 2 | -1.8 (0.2) | 0.2 (0.9) | <.001* | 2 | 1.6 (0.1) | 0.5 (0.8) | <.001* | 1 | -1.8 (0.2) | -0.2 (0.8) | <.001* |
|  | Beta | 2 | 1.7 (0.3) | -0.3 (1.1) | .132 | 2 | -2.0 (0.4) | 0.5 (0.8) | <.001* | 2 | 1.9 (0.2) | 0.9 (2.0) | .327 | 2 | -2.2 (0.5) | 0.5 (0.9) | <.001* |
|  | High Beta | 2 | 1.9 (0.2) | 1.7 (1.7) | .775 | 2 | -1.8 (0.2) | -0.9 (0.3) | <.001* | 2 | 1.9 (0.4) | 0.8 (0.7) | <.001* | 1 | -2.0 (0.3) | -0.5 (0.4) | <.001* |
|  | Beta 1 | 1 | 2.3 (0.6) | -0.6 (0.6) | <.001* | 2 | -1.8 (0.3) | -1.0 (0.2) | <.001* | 3 | 1.9 (0.4) | -0.5 (0.9) | <.001* | 2 | -1.8 (0.2) | -0.1 (0.7) | <.001* |
|  | Beta 2 | 1 | 1.6 (0.1) | -0.9 (0.1) | .003* | 2 | -1.7 (0.1) | -0.7 (0.2) | .001* | 2 | 2.0 (0.4) | 0.1 (0.3) | .004* | 2 | -2.1 (0.7) | -0.0 (0.6) | <.001* |
|  | Beta 3 | 2 | 1.7 (0.2) | 2.0 (1.3) | .655 | 2 | -1.9 (0.3) | -0.9 (0.3) | <.001* | 2 | 1.8 (0.2) | 0.6 (0.8) | <.001* | 2 | -2.2 (0.3) | -0.6 (0.4) | <.001* |
| Coherence | Delta | - | - | - | - | 8 | -2.6 (1.1) | -2.3 (1.5) | .029* | 3 | 2.3 (0.6) | -0.2 (1.1) | <.001* | 7 | -2.8 (1.2) | -2.3 (1.5) | <.001* |
|  | Theta | 5 | 2.0 (0.5) | -0.7 (1.0) | <.001* | 7 | -3.2 (1.6) | -2.8 (1.5) | .041* | 4 | 2.3 (0.7) | -0.2 (1.0) | <.001* | 7 | -3.1 (1.6) | -2.1 (1.5) | <.001* |
|  | Alpha | 5 | 1.9 (0.3) | -0.9 (1.2) | <.001* | 8 | -2.4 (0.9) | -2.4 (1.7) | .822 | 7 | 1.8 (0.3) | -0.3 (1.1) | <.001* | 7 | -3.0 (1.8) | -2.4 (1.7) | .052 |
|  | Beta | 6 | 2.3 (0.7) | -0.3 (1.1) | <.001* | 7 | -2.1 (0.5) | -2.4 (1.8) | .359 | 5 | 2.1 (0.5) | -0.2 (1.2) | <.001* | 7 | -2.4 (1.2) | -2.6 (1.6) | .420 |
| Asymmetry | Delta | 8 | 2.5 (0.9) | 0.4 (2.1) | <.001* | 8 | -2.4 (0.9) | -1.4 (1.8) | <.001* | 8 | 2.8 (1.1) | 0.4 (2.1) | <.001* | 7 | -2.8 (1.5) | -0.6 (1.3) | <.001* |
|  | Theta | 6 | 2.3 (1.0) | 0.4 (2.1) | <.001* | 8 | -2.3 (0.8) | -1.5 (1.6) | <.001* | 7 | 2.8 (1.5) | 0.1 (1.8) | <.001* | 8 | -2.6 (1.6) | -0.5 (1.6) | <.001* |
|  | Alpha | 3 | 2.3 (1.0) | 0.3 (1.3) | .096 | 5 | -2.0 (0.5) | -1.3 (1.7) | .025* | 4 | 3.3 (1.6) | -0.5 (2.2) | <.001* | 6 | -2.3 (1.4) | -0.2 (1.2) | <.001* |
|  | Beta | 5 | 1.7 (0.2) | -1.2 (1.3) | <.001* | 8 | -2.0 (0.5) | -0.8 (1.6) | <.001* | 5 | 2.5 (1.2) | -0.5 (2.0) | <.001* | 7 | -2.3 (0.9) | -0.7 (1.8) | <.001* |

After 6 months, the results for the delta wave (2–3.5 Hz) z-scores, showed that at least 3/8 subjects demonstrated a statistically significant decrease in absolute and relative powers towards normative values. When analyzing the coherence and asymmetry in this frequency band, all subjects had evidence of statistically significantly decreased absolute z-scores towards normative range in delta activity at 6 months. Theta wave (4–7.5 Hz) z-scores, showed a statistically significant decrease in absolute and relative powers in 3/8 subjects towards normative range; 7/8 subjects demonstrated decreases in the absolute z-score for coherence and all subjects demonstrated decreased absolute z-scores when assessing the asymmetry of this frequency band. Alpha wave (8–12.5 Hz) results revealed 2/8 subjects had significantly decreased absolute z-scores for absolute and relative power towards normative range; all subjects demonstrated evidence of decreased coherence in this frequency band with 6/8 subjects demonstrating decreased asymmetry absolute z-scores at 6 months. Low beta wave (13–20 Hz) activity in 2/8 subjects was significantly lower by absolute power and in 3/8 subjects by relative power (eyes open). A similar pattern was observed with high beta wave (20.5-28 Hz) activity.

### Activity Changes in Brodmann Areas

QEEG activity was mapped to Brodmann Areas of the brain using LORETA analysis with Brodman Area activity shown by the location of chronic pain, **Figure 1**. All the z-scores from the different Brodmann Areas were generated by the LORETA neurofeedback software and can be found for each arousal state and for each subject in **Supplementary Tables 2-9**. The clinically significant z-scores by channel and Brodmann Area for each subject individually can be found in **Supplementary Table 10**. Pre-treatment, Brodmann area activity was observed in the prefrontal cortex of the frontal lobe in Brodmann Areas 4 and 9 regardless of the location of pain, Figure 1. At 6 months, clinically significant activity in BAs 4 and 9 disappeared; statistically significant enhanced high beta wave activity was observed in Brodmann areas (BA) 21 (p=0.01, superolateral cortex of the temporal lobe), BA 29 and 30 (both retro splenial cortex of the limbic lobe p=0.04 and p=0.048, respectively), as shown in **Table 5**. In addition, increased clinically significant activity in the high beta waves in BA 19 (lateral occipital gyrus of the occipital lobe) and 39 (angular gyrus of the superolateral parietal lobe) approached significance (p=0.06).

**Table 5.** Changes in Brodmann Areas following focused muscle therapy evaluated with Eyes Closed and Eyes Open.

|  |  | EYES CLOSED |  |  |  | EYES OPEN |  |  |  |
| --- | --- | --- | --- | --- | --- | --- | --- | --- | --- |
| Freq. | Channels | Brodmann Area | Significance PRE (incidence) | Significance POST (incidence) | P-value (incidence) | Brodmann Area | Significance PRE (incidence) | Significance POST (incidence) | P-value (incidence) |
| Delta | 1 | 6 | 2.3 ± 0.9 (N=3) | - (N=0) | 0.06 | - | - | - | - |
| Theta | 5 | 40 | - (N=0) | 4.2 ± 0.2 (N=3) | 0.06 | - | - | - | - |
|  | 7 | - | - | - | - | 4 | 2.8 ± 0.2 (N=3) | - (N=0) | 0.06 |
| Alpha | 11 | - | - | - | - | 9 | 2.6 ± 1.0 (N=4) | - (N=0) | 0.02* |
| Beta | 15 | 9 | 2.6 ± 0.5 (N=3) | - (N=0) | 0.06 | - | - | - | - |
| High Beta | 19 | 8 | 2.1 ± 0.6 (N=3) | - (N=0) | 0.06 | 19 | - (N=0) | 2.1 ± 0.7 (N=3) | 0.06 |
|  | 21 | 40 | - (N=0) | 1.8 ± 0.9 (N=5) | <.01* | 39 | - (N=0) | 2.0 ± 0.9 (N=3) | 0.06 |
|  | 22 | 7 | - (N=0) | 1.9 ± 1.4 (N=3) | 0.06 | - | - | - | - |
|  | 23 | 8 | 1.6 ± 0.3 (N=3) | 0 (N=0) | 0.06 | - | - | - | - |
|  | 24 | 8 | 1.6 ± 0.3 (N=3) | - (N=0) | 0.06 | - | - | - | - |
|  | 29 | 7 | - (N=0) | 2.0 ± 0.8 (N=5) | 0.04* | - | - | - | - |
|  | 30 | 7 | 2.1 ± 0.2 (N=2) | 2.2 ± 0.9 (N=6) | 0.05* | - | - | - | - |

**Figure 1.**
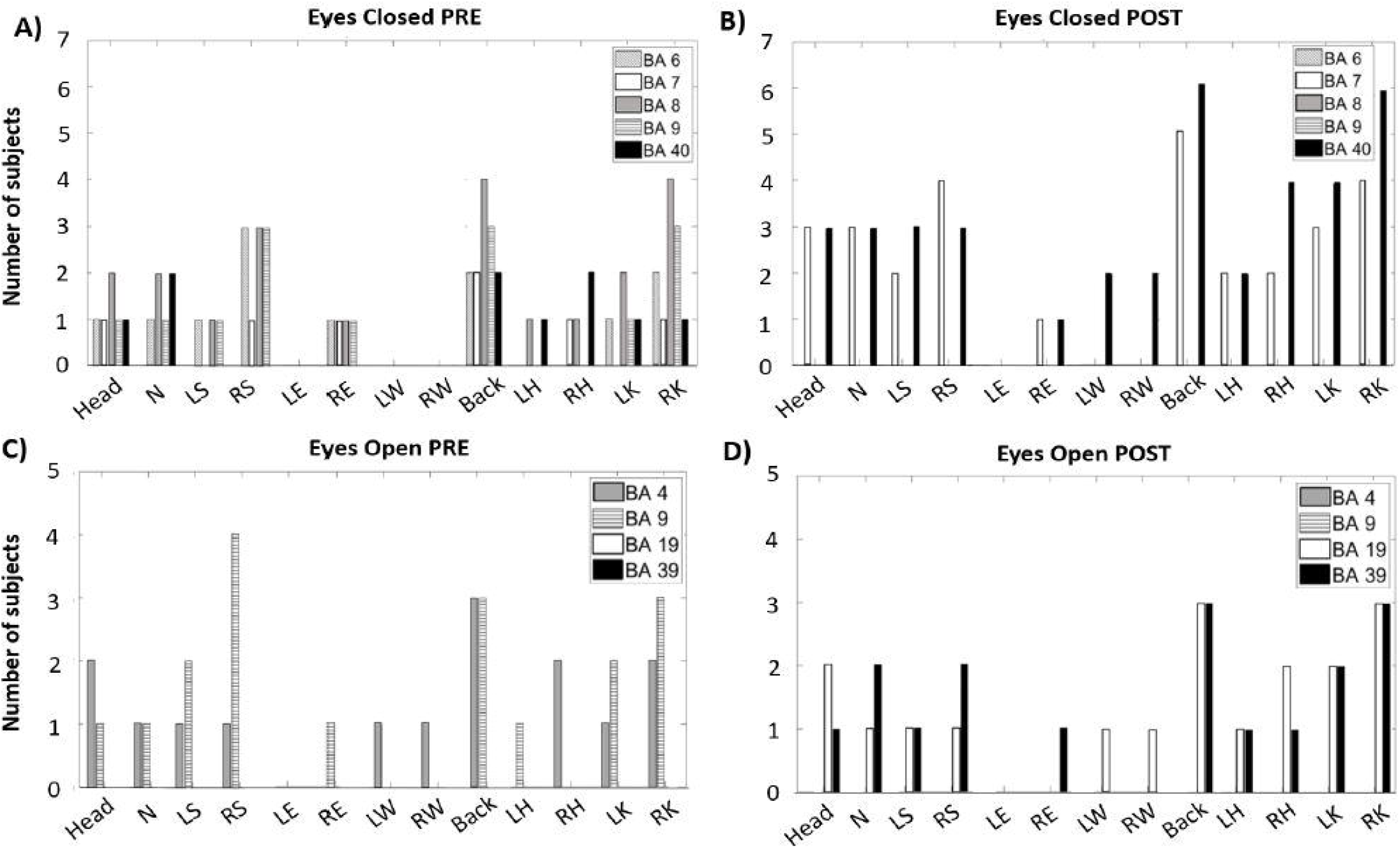
Brodmann Areas of interest by pain location. *{Head, N-neck, LS-left shoulder, RS-right shoulder, LE-left elbow, RE-right elbow, LW-left wrist, RW-right wrist, Back, LH-left hip, RH-right hip, LK-left knee, RK-right knee*

### Analyses of auditory and visual attention

The IVA-2 Standard Full-Scale analysis was used to measure visual and auditory attentiveness. At 6 months post-treatment, there were clinically relevant improvements in attention, **Table 6**; mean auditory attention increased from slightly impaired (IVA score 87.5 ± 28.8) to average (IVA score 99.6 ± 22.1). Additional clinically relevant improvement was noted in visual stamina which improved from mildly impaired, 83.5±15.5 to average 90.4±17.0

**Table 6.** IVA-2 Standard Scale Analysis.

|  | FULL-SCALE |  |  |  | AUDITORY |  |  |  | VISUAL |  |  |  |
| --- | --- | --- | --- | --- | --- | --- | --- | --- | --- | --- | --- | --- |
| Parameter | PRE |  | POST |  | PRE |  | POST |  | PRE |  | POST |  |
|  | Mean (SD) | Range of Mean | Mean (SD) | Range of Mean | Mean (SD) | Range of Mean | Mean (SD) | Range of Mean | Mean (SD) | Range of Mean | Mean (SD) | Range of Mean |
| Attention Quotient | 94.0 (18.4) | Average | 95.8 (18.2) | Average | 91.0 (19.6) | Average | 98.6 (16.6) | Average | 98.5 (16.7) | Average | 94.4 (21.5) | Average |
| Sustained Attention |  |  |  |  | 87.5 (28.8) | Slightly impaired | 99.6 (22.1) | Average* | 97.6 (21.5) | Average | 94.8 (22.8) | Average |
| Vigilance |  |  |  |  | 85.5 (32.7) | Slightly impaired | 95.9 (15.9) | Average | 102.6 (10.2) | Average | 95.9 (18.1) | Average |
| Focus |  |  |  |  | 91.0 (12.0) | Average | 96.4 (13.8) | Average | 98.5 (17.9) | Average | 98.5 (18.3) | Average |
| Speed |  |  |  |  | 103.7 (5.4) | Average | 104.3 (11.4) | Average | 95.5 (9.4) | Average | 93.9 (14.2) | Average |
| Response Control Quotient | 89.6 (16.0) | Slightly impaired | 99.8 (19.6) | Average | 94.9 (14.4) | Average | 99.8 (16.7) | Average | 92.1 (16.0) | Average | 99.9 (16.5) | Average |
| Prudence |  |  |  |  | 105.4 (17.9) | Average | 107.1 (10.5) | Average | 99.6 (14.6) | Average | 108.3 (8.9) | Average |
| Consistency |  |  |  |  | 88.6 (14.3) | Slightly impaired | 99.8 (15.5) | Average | 98.9 (18.1) | Average | 102.0 (12.3) | Average |
| Stamina |  |  |  |  | 91.5 (17.3) | Average | 92.9 (13.3) | Average | 83.5 (15.5) | Mildly impaired | 90.4 (17.0) | Average* |
| Comprehension |  |  |  |  | 97.0 (19.3) | Average | 102.9 (10.0) | Average | 93.9 (20.0) | Average | 101.0 (9.5) | Average |
| Persistence |  |  |  |  | 94.6 (17.3) | Average | 82.8 (9.4) | Mildly impaired | 104.4 (25.2) | Average | 93.5 (11.2) | Average |
| Sensory/Motor |  |  |  |  | 112.6 (4.5) | Above average | 110.5 (9.3) | Above average | 95.8 (17.6) | Average | 104.8 (8.5) | Average |

**Ranges legend:** X ≤ 60 (Extremely impaired); [61 – 67] (Severely impaired); [68 – 71] (Moderately – Severely impaired); [72 – 75] (Moderately impaired); [76 – 79] (Mildly – Moderately impaired); [80 – 84] (Mildly impaired); [85 – 89] (Slightly impaired); [90 – 109] (Average); [110 – 119] (Above Average); [120 – 129] (Superior); X ≥ 130 (Exceptional).
\* Change in Parameter from Pre to Post was clinically significant

## DISCUSSION

Retired NFL players demonstrated multiple sites of chronic pain; the pain was present for a minimum of 5 years and persisted despite multiple therapies. Based on our previous report on renal transplant recipients [22] in which subjects reported significant pain relief with therapy, we hypothesized that similar pain relief could be achieved in retired NFL players, who are known to have chronic pain following multiple traumatic injuries on the field. Our results indicated that focused muscle contraction therapy led to the cessation or decrease of chronic pain and the cessation or decrease of pain was associated with significant changes in brain wave activity.

In patients with chronic pain, brain wave activity has most commonly increased in the theta bandwidth [7; 9; 39]. It is believed that increased theta wave activity is associated with thalamocortical dysrhythmia, creating thalamocortical loops which may lead to abnormal gamma wave activity and the symptoms of chronic pain [12; 27]. It is also believed that increased theta wave activity in the awake patient with pain, is associated with slowing of EEG activity and in some cases, sleepiness, or decreased attentiveness, since theta waves are primarily observed in dream-like states [24]. Chronic pain has also been reported to associate with increased activity in alpha and beta waves, particularly in the primary somatosensory and medial prefrontal cortex with beta waves likely to be observed in the frontal brain areas [21]. The results shown in this study are consistent with reports of others; we observed Z-scores outside normative ranges in delta, theta, and beta wave activities pre-therapy in all subjects with subsequent return to normative ranges at 6 months post-therapy. Importantly, these changes were associated with the cessation of chronic pain.

The location of brain wave activity was observed in the prefrontal cortex of the frontal lobe in Brodman Areas 4 and 9; these findings are consistent with prior reports of brain areas associated with chronic pain [28; 35]. Interestingly, these two areas showed activity regardless of the location of the pain. At 6 months post-treatment, none of the subjects showed clinically significant activity in BA 4 and 9. Instead, statistically significant activity was observed in BA 21, 29, and 30. BA 21 is located in the temporal cortex and is responsible for auditory processing and language [1]. BA 29 and 30 are located in the retro splenial regions of the cerebral cortex (RSC)[31]. RSC activation occurs during spatial awareness for activities such as route planning, trajectory changes, thoughts of location, and orientation. Overall fMRI studies point to its role in both encoding and retrieval of spatial information.

An increased activity in the high beta waves of BA 39 (angular gyrus of the superolateral parietal lobe) which has played a role in the patient’s own-body perceptions [15], approached statistical significance. Body perceptions are known to influence the perception of chronic pain [3]. The increased activity in the high beta waves of BA 19 (lateral occipital gyrus of the occipital lobe) was also observed to approach statistical significance. BA 19 has been identified as part of the visual cortex and is involved in the representation and perception of objects [29]. This area is strongly activated when objects are touched and plays a role in behavioral performance in different recognition tasks and response to a variety of types of stimuli [18].

Improvements in auditory attention and visual stamina may be due to the significantly increased brain activity in high beta waves observed in BA 21 and BA 39, respectively. High beta waves are the highest form of mental alertness. The specific task of focusing on isolated contraction of singular muscle groups during therapy may involve the temporal-parietal-occipital (TPO) junction, a complex area of the brain which is involved in several high-level neurological functions, such as language, visuospatial recognition, self-processing- and working memory. This area includes BAs19, 37, 39, and 40; we observed increased activity in two of these, BA 19 and 39. Focused muscle contraction relies on auditory coaching during therapy which could account for activation of language function. Visuospatial recognition is used in visualizing muscle contraction during repetitions. Self-processing is applied in response to feedback by the therapist to guide the patient on the accuracy of focused muscle contraction. Working memory is used to recall appropriate techniques used for precise isolation of the muscle group. In summary, focused muscle contraction through its execution, engaged a variety of tasks requiring concentration and focused mental activity.

This study had limitations. The selection of retired NFL players, who by nature have the highest rate of injury and as a consequence, a high number of locations manifesting chronic pain, did inherently limit the number of subjects enrolled at this single-site study. A randomized clinical trial is theoretically needed to confirm the validity of this pilot study. Because of the great chronicity of the pain (>5 years) and the numerous unsuccessful treatment therapies during that period, randomizing a treatment arm to no therapy would likely prolong the chronic pain condition and may not be considered compassionate care. Further, this is the second study in an entirely different population, retired NFL players, which has led to consistent relief of pain; the first study, a randomized control trial, was undertaken in 80 renal transplant recipients with similar results using the analog scale of pain. We believe this non-medication-based therapy engendered a high motivation to continue therapy to 18 months and beyond due to substantive reduction or elimination of chronic pain. This program demonstrates feasibility as an effective and long-lasting treatment for patients suffering from chronic pain and its detriments.

## Data Availability

All data produced in the present study are available upon reasonable request to the authors.

## Acknowledgments

This study was supported by a grant from the Department of Surgery and the University of Illinois Foundation. GH, MH, DZ are employed by GH Fitlab but performed muscle therapy in collaboration with the College of Kinesiology at the University of Illinois. NC and MO are employed by Holistic Neuropsychology but performed all neuropsychology assessments at the University of Illinois. No other conflict of interests is acknowledged.

