## Supplementary Table for "The effect of focused muscle contraction therapy on chronic pain and Brodmann Area activity in former National Football League players"

**Supplementary Table 1. Focused Muscle Contraction Initial weights by Subject and muscle group**

| Body site | Ultra-Low Starting Weights with NFL Football players | | | | | | | |
|---|---|---|---|---|---|---|---|---|
| | S1 | S2 | S3 | S4 | S5 | S6 | S7 | S8 |
| **R. Anterior tibialis** | 7 | 10 | 38 | 40 | 40 | 7 | 10 | 5 |
| **L. Anterior tibialis** | 7 | 10 | 40 | 40 | 40 | 7 | 10 | 5 |
| **Low Row** | 17 | 18.5 | 30 | 22 | 40 | 17.5 | 20 | 20 |
| **Gluteus** | 20 | 22.5 | 30 | 25 | 45 | 17.5 | 22 | 20 |
| **Pulldown** | 25 | 18.5 | 40 | 25 | 40 | 17.5 | 20 | 17.5 |
| **Hamstring** | 20 | 18.5 | 30 | 22 | 40 | 17.5 | 20 | 20 |
| **Leg Extension.** | 10 | 10 | 22 | 10 | 15 | 5 | 10 | 7.5 |
| **Biceps** | 33 | 22.5 | 20 | 20 | 30 | 20 | 25 | 20 |
| **Triceps** | 20 | 15 | 18 | 20 | 20 | 16 | 18 | 20 |
| **R. Calf** | 10 | 10 | 25 | 35 | 30 | 10 | 15 | 10 |
| **L. Calf** | 10 | 10 | 25 | 35 | 30 | 10 | 15 | 10 |
| **MEAN** | 16.3 | 15.0 | 28.9 | 26.7 | 33.6 | 13.2 | 16.8 | 14.1 |

**Supplemental Table 2. Z-scores by Brodmann Area by arousal state for Subject 1 (S1)**

| S1 | | Eyes Closed | | | | | | Eyes Open | | | | | |
|---|---|---|---|---|---|---|---|---|---|---|---|---|---|
| | | PRE | | | POST | | | PRE | | | Post | | |
| Arousal State | Freq. (Hz) | Side | Brodmann Area | Z-score | Side | Brodmann Area | Z-score | Side | Brodmann Area | Z-score | Side | Brodmann Area | Z-score |
| Delta (deep sleep) | 1 | L | 6 | 2.3 | R | 37 | 3.5 | L | 13 | 3.15 | L | 13 | 3.3 |
| Delta (deep sleep) | 2 | R | 2 | 2.4 | R | 37 | 2.8 | L | 13 | 3.8 | L | 18 | 2.8 |
| Delta (deep sleep) | 3 | R | 40 | 2.7 | R | 4 | 2.5 | L | 13 | 3.9 | L | 18 | 3 |
| Delta (deep sleep) | 4 | R | 40 | 3.5 | R | 4 | 3 | L | 40 | 3.6 | L | 19 | 2.9 |
| theta(drowsy) | 5 | R | 9 | 3.2 | R | 1 | 2.7 | L | 40 | 3.6 | L | 18 | 3 |
| theta(drowsy) | 6 | R | 9 | 3.5 | R | 4 | 2.6 | L | 4 | 3.8 | R | 18 | 2.9 |
| theta(drowsy) | 7 | R | 8 | 3.3 | L | 38 | 2.5 | L | 4 | 2.7 | L | 18 | 3 |
| alpha(wakeful) | 8 | R | 9 | 3 | R | 6 | 1.9 | R | 8 | 2.4 | L | 18 | 2.3 |
| alpha(wakeful) | 9 | R | 9 | 3.5 | L | 9 | 1.4 | R | 8 | 2.7 | L | 19 | 1.5 |
| alpha(wakeful) | 10 | R | 9 | 2.8 | L | 9 | 1.3 | L | 8 | 2.8 | L | 19 | 1.2 |
| alpha(wakeful) | 11 | R | 9 | 2.8 | L | 9 | 1.2 | L | 9 | 3.1 | L | 19 | 1.5 |
| beta (relaxed alert) | 12 | R | 9 | 3 | L | 9 | 1.2 | L | 9 | 2.7 | L | 18 | 2.2 |
| beta (relaxed alert) | 13 | R | 9 | 2.9 | L | 6 | 1.3 | L | 9 | 3 | L | 18 | 2.4 |
| beta (relaxed alert) | 14 | R | 9 | 2.4 | L | 6 | 1.2 | R | 9 | 2.4 | L | 18 | 2.4 |
| beta (relaxed alert) | 15 | R | 9 | 2.3 | R | 40 | 1.1 | L | 17 | 2 | L | 19 | 2.4 |
| beta (relaxed alert) | 16 | R | 8 | 2.1 | R | 40 | 0.9 | L | 17 | 1.6 | L | 18 | 2.1 |
| beta (relaxed alert) | 17 | R | 9 | 2 | L | 40 | 0.8 | R | 9 | 1.3 | L | 18 | 2.1 |
| beta (relaxed alert) | 18 | R | 9 | 2 | R | 40 | 0.8 | L | 18 | 1.2 | L | 19 | 1.6 |
| high beta (aroused and actively engaged) | 19 | R | 9 | 1.6 | R | 40 | 0.8 | R | 9 | 1 | L | 19 | 1.4 |
| high beta (aroused and actively engaged) | 20 | R | 9 | 1.4 | R | 40 | 0.7 | L | 40 | 0.9 | L | 19 | 1.3 |
| high beta (aroused and actively engaged) | 21 | R | 9 | 1.4 | R | 40 | 0.9 | L | 18 | 1.2 | L | 19 | 1.6 |
| high beta (aroused and actively engaged) | 22 | R | 8 | 1.4 | L | 7 | 0.9 | R | 18 | 1.3 | L | 18 | 1.9 |
| high beta (aroused and actively engaged) | 23 | R | 8 | 1.6 | L | 7 | 1 | L | 18 | 1 | L | 18 | 2 |
| high beta (aroused and actively engaged) | 24 | R | 8 | 1.4 | R | 40 | 1.1 | L | 18 | 1.3 | L | 18 | 1.7 |
| high beta (aroused and actively engaged) | 25 | R | 9 | 1.4 | L | 7 | 1.1 | L | 10 | -1 | R | 13 | -1.3 |
| high beta (aroused and actively engaged) | 26 | R | 8 | 1.3 | L | 7 | 1.3 | L | 18 | 1 | R | 13 | -1.4 |
| high beta (aroused and actively engaged) | 27 | R | 9 | 1.4 | L | 7 | 1.3 | L | 32 | -1 | R | 13 | -1.4 |
| high beta (aroused and actively engaged) | 28 | R | 9 | 1.4 | L | 7 | 1.4 | L | 10 | -1.1 | R | 18 | 1.4 |
| high beta (aroused and actively engaged) | 29 | R | 9 | 1.3 | L | 7 | 1.3 | R | 18 | 1.2 | R | 18 | 1.5 |
| high beta (aroused and actively engaged) | 30 | R | 40 | 1.5 | R | 40 | 1.2 | R | 18 | 1.3 | R | 18 | 1.6 |

### Supplemental Table 3. Z-scores by Brodmann Area by arousal state for Subject 2 (S2)

| S2 | | Eyes Closed | | | | | | Eyes Open | | | | | |
|---|---|---|---|---|---|---|---|---|---|---|---|---|---|
| | | PRE | | | POST | | | PRE | | | Post | | |
| Arousal State | Freq. (Hz) | Side | Brodmann Area | Z-score | Side | Brodmann Area | Z-score | Side | Brodmann Area | Z-score | Side | Brodmann Area | Z-score |
| Delta (deep sleep) | 1 | R | 32 | 2.8 | L | 13 | 2.8 | L | 31 | 3.4 | R | 13 | 2.1 |
| Delta (deep sleep) | 2 | L | 24 | 3 | R | 13 | 3.2 | L | 31 | 3.4 | R | 13 | 3.1 |
| Delta (deep sleep) | 3 | R | 24 | 3.3 | R | 13 | 4 | R | 29 | 3.6 | R | 13 | 3.7 |
| Delta (deep sleep) | 4 | R | 40 | 3.4 | R | 13 | 3.9 | R | 41 | 4.5 | R | 13 | 3.6 |
| theta(drowsy) | 5 | L | 11 | 3.7 | R | 40 | 4 | R | 42 | 4.6 | R | 40 | 4 |
| theta(drowsy) | 6 | L | 11 | 3.3 | R | 47 | 4.3 | R | 21 | 4.2 | R | 40 | 4 |
| theta(drowsy) | 7 | L | 11 | 2.9 | R | 47 | 3.9 | R | 21 | 3.5 | R | 9 | 3.8 |
| alpha(wakeful) | 8 | R | 8 | 2.6 | R | 8 | 3.1 | R | 21 | 3.2 | R | 8 | 3.1 |
| alpha(wakeful) | 9 | R | 9 | 2.8 | R | 47 | 2.6 | R | 9 | 2.6 | L | 9 | 2.8 |
| alpha(wakeful) | 10 | R | 8 | 3 | R | 9 | 2.7 | R | 8 | 2 | L | 6 | 3.3 |
| alpha(wakeful) | 11 | L | 9 | 3 | R | 21 | 2.9 | R | 9 | 2.4 | L | 8 | 3.2 |
| beta (relaxed alert) | 12 | R | 9 | 3.3 | L | 9 | 2.8 | R | 8 | 2.5 | L | 10 | 3.3 |
| beta (relaxed alert) | 13 | R | 9 | 3 | L | 6 | 2.7 | R | 21 | 2.3 | R | 6 | 3.3 |
| beta (relaxed alert) | 14 | R | 8 | 2.6 | L | 6 | 2.4 | R | 21 | 2.3 | L | 6 | 2.9 |
| beta (relaxed alert) | 15 | R | 9 | 2.3 | R | 6 | 2.2 | R | 21 | 1.9 | L | 1 | 2.6 |
| beta (relaxed alert) | 16 | R | 9 | 1.9 | R | 40 | 2 | R | 21 | 2.1 | L | 1 | 2.5 |
| beta (relaxed alert) | 17 | R | 8 | 1.8 | L | 6 | 1.8 | R | 20 | 2.2 | L | 4 | 2.3 |
| beta (relaxed alert) | 18 | R | 8 | 1.8 | R | 40 | 1.7 | R | 9 | 1.9 | L | 4 | 2.2 |
| high beta (aroused and actively engaged) | 19 | R | 8 | 1.5 | R | 40 | 1.6 | L | 11 | 1.6 | L | 37 | 1.9 |
| high beta (aroused and actively engaged) | 20 | R | 8 | 1.5 | R | 40 | 1.6 | L | 11 | 1.8 | L | 6 | 1.8 |
| high beta (aroused and actively engaged) | 21 | R | 8 | 1.3 | R | 40 | 1.8 | L | 11 | 1.7 | L | 39 | 1.8 |
| high beta (aroused and actively engaged) | 22 | R | 8 | 1.3 | R | 40 | 1.6 | R | 7 | 1.6 | L | 40 | 1.7 |
| high beta (aroused and actively engaged) | 23 | R | 8 | 1.3 | R | 40 | 1.7 | R | 21 | 1.3 | L | 22 | 2.2 |
| high beta (aroused and actively engaged) | 24 | R | 8 | 1.3 | R | 40 | 1.7 | L | 7 | 1.7 | R | 39 | 1.6 |
| high beta (aroused and actively engaged) | 25 | R | 7 | 1.3 | R | 39 | 1.5 | R | 18 | 1.7 | R | 40 | 1.5 |
| high beta (aroused and actively engaged) | 26 | R | 7 | 1.3 | R | 40 | 1.7 | L | 18 | 1.8 | R | 40 | 1.3 |
| high beta (aroused and actively engaged) | 27 | L | 40 | 1.4 | R | 40 | 1.4 | L | 7 | 1.7 | L | 7 | 1.5 |
| high beta (aroused and actively engaged) | 28 | L | 40 | 1.6 | R | 7 | 1.6 | L | 7 | 2 | L | 40 | 1.5 |
| high beta (aroused and actively engaged) | 29 | L | 40 | 1.6 | R | 39 | 1.4 | R | 40 | 1.8 | L | 40 | 1.6 |
| high beta (aroused and actively engaged) | 30 | L | 40 | 1.8 | R | 7 | 1.5 | L | 40 | 2 | L | 40 | 1.9 |

**Supplemental Table 4. Z-scores by Brodmann Area by arousal state for Subject 3 (S3)**

| S3 | | Eyes Closed | | | | | | Eyes Open | | | | | |
|---|---|---|---|---|---|---|---|---|---|---|---|---|---|
| | | PRE | | | POST | | | PRE | | | Post | | |
| Arousal State | Freq. (Hz) | Side | Brodmann Area | Z-score | Side | Brodmann Area | Z-score | Side | Brodmann Area | Z-score | Side | Brodmann Area | Z-score |
| Delta (deep sleep) | 1 | R | 6 | 1.3 | R | 9 | 2.1 | R | 4 | 1.1 | R | 22 | 3.4 |
| Delta (deep sleep) | 2 | R | 4 | 2 | L | 6 | 2.3 | R | 4 | 1.7 | R | 41 | 3.3 |
| Delta (deep sleep) | 3 | R | 4 | 2.4 | R | 8 | 2.8 | R | 4 | 2.3 | L | 6 | 3.9 |
| Delta (deep sleep) | 4 | R | 4 | 2.8 | L | 6 | 3.1 | R | 4 | 2.6 | R | 41 | 4.1 |
| theta(drowsy) | 5 | R | 4 | 2.7 | R | 9 | 3.2 | R | 4 | 2.6 | R | 41 | 4.3 |
| theta(drowsy) | 6 | R | 4 | 2.6 | R | 9 | 3.5 | R | 4 | 2.7 | R | 22 | 4.6 |
| theta(drowsy) | 7 | R | 4 | 2.1 | R | 9 | 3.9 | R | 9 | 2.5 | R | 10 | 4.7 |
| alpha(wakeful) | 8 | R | 6 | 1.5 | R | 9 | 3.3 | R | 9 | 1.7 | R | 8 | 4.1 |
| alpha(wakeful) | 9 | R | 9 | 1 | R | 9 | 3.2 | R | 9 | 1.5 | R | 9 | 3.4 |
| alpha(wakeful) | 10 | R | 9 | 1 | L | 8 | 3.2 | L | 8 | 1.3 | L | 8 | 2.6 |
| alpha(wakeful) | 11 | R | 9 | 0.8 | R | 6 | 3.1 | R | 9 | 1.4 | R | 6 | 2.8 |
| beta (relaxed alert) | 12 | R | 9 | 1 | L | 9 | 3.5 | R | 4 | 1.5 | L | 9 | 3.4 |
| beta (relaxed alert) | 13 | R | 9 | 1.3 | R | 9 | 3.3 | R | 4 | 1.9 | R | 6 | 3.1 |
| beta (relaxed alert) | 14 | R | 40 | 1.4 | L | 8 | 3 | L | 6 | 1.7 | L | 9 | 2.6 |
| beta (relaxed alert) | 15 | R | 40 | 1.4 | L | 8 | 2.6 | R | 40 | 1.5 | L | 8 | 2.4 |
| beta (relaxed alert) | 16 | R | 40 | 1.5 | L | 8 | 2.3 | R | 40 | 1.3 | R | 6 | 2.1 |
| beta (relaxed alert) | 17 | R | 21 | 1.5 | R | 8 | 2 | R | 21 | 1.2 | R | 6 | 2 |
| beta (relaxed alert) | 18 | R | 21 | 1.6 | R | 6 | 1.8 | R | 21 | 1.2 | R | 6 | 2 |
| high beta (aroused and actively engaged) | 19 | R | 11 | 1.6 | R | 6 | 1.5 | R | 21 | 1.5 | R | 6 | 2 |
| high beta (aroused and actively engaged) | 20 | R | 11 | 1.6 | R | 9 | 1.5 | R | 40 | 1.2 | R | 8 | 2.1 |
| high beta (aroused and actively engaged) | 21 | R | 11 | 1.6 | R | 9 | 1.5 | R | 37 | 1.2 | R | 9 | 1.5 |
| high beta (aroused and actively engaged) | 22 | R | 11 | 1.6 | R | 8 | 1.3 | R | 20 | 1.1 | R | 9 | 1.6 |
| high beta (aroused and actively engaged) | 23 | R | 11 | 1.4 | R | 9 | 1.2 | R | 21 | 1.2 | R | 9 | 1.2 |
| high beta (aroused and actively engaged) | 24 | R | 11 | 1.2 | R | 9 | 1.2 | R | 20 | 1.3 | R | 9 | 1.2 |
| high beta (aroused and actively engaged) | 25 | R | 39 | 1.2 | R | 9 | 1.1 | R | 20 | 1.4 | R | 6 | 1 |
| high beta (aroused and actively engaged) | 26 | R | 39 | 1.3 | R | 9 | 1.1 | R | 20 | 1.3 | R | 7 | 1.1 |
| high beta (aroused and actively engaged) | 27 | R | 7 | 1.5 | R | 9 | 1.1 | R | 38 | 1.3 | R | 9 | 0.9 |
| high beta (aroused and actively engaged) | 28 | R | 7 | 1.4 | R | 6 | 1.2 | R | 20 | 1.1 | R | 7 | 1.5 |
| high beta (aroused and actively engaged) | 29 | R | 40 | 1.3 | R | 7 | 1.4 | R | 40 | 1.1 | R | 32 | 1.4 |
| high beta (aroused and actively engaged) | 30 | R | 40 | 1.4 | R | 7 | 1.2 | R | 40 | 1.2 | R | 9 | 1.1 |

**Supplemental Table 5. Z-scores by Brodmann Area by arousal state for Subject 4 (S4)**

| **S4** | | Eyes Closed | | | | | | Eyes Open | | | | | |
|---|---|---|---|---|---|---|---|---|---|---|---|---|---|
| | | PRE | | | POST | | | PRE | | | Post | | |
| **Arousal State** | **Freq. (Hz)** | **Side** | **Brodmann Area** | **Z-score** | **Side** | **Brodmann Area** | **Z-score** | **Side** | **Brodmann Area** | **Z-score** | **Side** | **Brodmann Area** | **Z-score** |
| Delta (deep sleep) | 1 | R | 13 | 2.4 | L | 13 | 2.5 | R | 13 | 2.4 | R | 9 | 1.3 |
| Delta (deep sleep) | 2 | R | 13 | 3 | L | 6 | 2.9 | R | 2 | 1.6 | L | 24 | 2.2 |
| Delta (deep sleep) | 3 | R | 13 | 3.1 | L | 6 | 3.6 | L | 4 | 2.3 | L | 6 | 3.1 |
| Delta (deep sleep) | 4 | R | 4 | 3.1 | L | 6 | 3.7 | L | 4 | 3.1 | R | 4 | 3.1 |
| theta(drowsy) | 5 | R | 2 | 2.8 | L | 4 | 4 | L | 4 | 3 | R | 4 | 3.2 |
| theta(drowsy) | 6 | R | 4 | 2.7 | L | 4 | 4.2 | L | 4 | 3.3 | L | 4 | 3.5 |
| theta(drowsy) | 7 | R | 3 | 2.5 | L | 4 | 3.6 | L | 4 | 3 | R | 9 | 3.5 |
| alpha(wakeful) | 8 | R | 9 | 2.2 | L | 6 | 2.6 | L | 8 | 2.5 | L | 10 | 3.4 |
| alpha(wakeful) | 9 | R | 9 | 1.7 | L | 20 | 2.1 | R | 8 | 2.3 | R | 8 | 2.9 |
| alpha(wakeful) | 10 | R | 9 | 1.3 | R | 8 | 1.7 | R | 9 | 1.9 | L | 8 | 2.7 |
| alpha(wakeful) | 11 | R | 9 | 1.4 | R | 8 | 1.8 | L | 10 | 1.4 | L | 8 | 2.5 |
| beta (relaxed alert) | 12 | R | 9 | 1.5 | R | 9 | 2 | L | 1 | 2.3 | R | 6 | 3.2 |
| beta (relaxed alert) | 13 | L | 6 | 1.7 | L | 1 | 2.6 | L | 40 | 2.7 | R | 6 | 3.6 |
| beta (relaxed alert) | 14 | R | 40 | 1.7 | L | 40 | 2.9 | L | 40 | 2.2 | R | 8 | 3.1 |
| beta (relaxed alert) | 15 | R | 40 | 1.7 | L | 40 | 2.8 | L | 40 | 1.9 | L | 8 | 2.8 |
| beta (relaxed alert) | 16 | R | 21 | 1.8 | L | 40 | 2.5 | L | 40 | 2.5 | R | 8 | 2.7 |
| beta (relaxed alert) | 17 | R | 40 | 1.9 | L | 40 | 2.3 | L | 5 | 1.6 | R | 6 | 2.7 |
| beta (relaxed alert) | 18 | L | 40 | 1.4 | L | 40 | 2.6 | L | 40 | 2.1 | R | 6 | 2.4 |
| high beta (aroused and actively engaged) | 19 | L | 7 | 1.4 | L | 40 | 2.2 | L | 40 | 1.8 | R | 8 | 2.1 |
| high beta (aroused and actively engaged) | 20 | L | 40 | 1.2 | L | 40 | 2.2 | L | 7 | 2.2 | R | 6 | 2.3 |
| high beta (aroused and actively engaged) | 21 | L | 40 | 1.3 | L | 40 | 2.4 | L | 7 | 1.6 | L | 7 | 2.9 |
| high beta (aroused and actively engaged) | 22 | L | 40 | 1.5 | L | 40 | 2.2 | R | 7 | 2.2 | R | 7 | 2.6 |
| high beta (aroused and actively engaged) | 23 | L | 7 | 1.8 | L | 40 | 2.6 | L | 7 | 1.9 | R | 7 | 2.6 |
| high beta (aroused and actively engaged) | 24 | R | 7 | 1.8 | L | 7 | 2.5 | L | 7 | 2.1 | L | 7 | 2.4 |
| high beta (aroused and actively engaged) | 25 | L | 7 | 2.2 | L | 7 | 2.6 | R | 7 | 2.1 | R | 7 | 2.9 |
| high beta (aroused and actively engaged) | 26 | L | 7 | 2.5 | L | 7 | 2.7 | R | 7 | 2 | R | 7 | 3 |
| high beta (aroused and actively engaged) | 27 | L | 7 | 2.4 | L | 7 | 2.9 | R | 7 | 2 | R | 7 | 2.8 |
| high beta (aroused and actively engaged) | 28 | L | 7 | 2.7 | L | 7 | 3.1 | R | 7 | 2 | R | 7 | 3 |
| high beta (aroused and actively engaged) | 29 | L | 7 | 2.2 | L | 7 | 2.7 | L | 7 | 2 | R | 7 | 2.7 |
| high beta (aroused and actively engaged) | 30 | L | 7 | 2.5 | L | 7 | 2.8 | R | 7 | 2.5 | R | 7 | 2.9 |

### Supplemental Table 6. Z-scores by Brodmann Area by arousal state for Subject 5 (S5)

| S5 | | Eyes Closed | | | | | | Eyes Open | | | | | |
|---|---|---|---|---|---|---|---|---|---|---|---|---|---|
| | | PRE | | | POST | | | PRE | | | Post | | |
| Arousal State | Freq. (Hz) | Side | Brodmann Area | Z-score | Side | Brodmann Area | Z-score | Side | Brodmann Area | Z-score | Side | Brodmann Area | Z-score |
| Delta (deep sleep) | 1 | R | 32 | 4 | L | 13 | 2.9 | R | 13 | 8.4 | R | 37 | 1.9 |
| Delta (deep sleep) | 2 | L | 23 | 5 | L | 13 | 3.2 | R | 13 | 8.1 | L | 19 | 2.6 |
| Delta (deep sleep) | 3 | L | 45 | 4.4 | L | 13 | 4 | R | 13 | 8.4 | R | 22 | 3.4 |
| Delta (deep sleep) | 4 | R | 13 | 48 | L | 40 | 3.9 | R | 29 | 8.1 | L | 9 | 3.5 |
| theta(drowsy) | 5 | L | 45 | 5.4 | L | 40 | 4.3 | R | 29 | 8.8 | L | 9 | 4 |
| theta(drowsy) | 6 | L | 45 | 5.4 | L | 46 | 4.4 | R | 44 | 9.6 | R | 9 | 3.6 |
| theta(drowsy) | 7 | L | 45 | 5.3 | L | 10 | 3.9 | R | 9 | 8.6 | R | 9 | 3.9 |
| alpha(wakeful) | 8 | L | 46 | 4.6 | L | 10 | 3.3 | R | 8 | 6.9 | R | 9 | 2.8 |
| alpha(wakeful) | 9 | R | 8 | 4.9 | L | 10 | 3.4 | R | 8 | 6.1 | R | 9 | 2.7 |
| alpha(wakeful) | 10 | R | 8 | 4.6 | L | 8 | 3.9 | R | 10 | 6 | L | 8 | 3.1 |
| alpha(wakeful) | 11 | R | 9 | 4.5 | R | 9 | 4.2 | R | 8 | 6 | L | 8 | 3.4 |
| beta (relaxed alert) | 12 | R | 8 | 4.2 | R | 9 | 4.2 | R | 6 | 7.1 | R | 8 | 2.9 |
| beta (relaxed alert) | 13 | R | 6 | 3.8 | R | 9 | 4.2 | R | 6 | 7.4 | R | 9 | 3.1 |
| beta (relaxed alert) | 14 | R | 6 | 3.8 | L | 8 | 3.6 | R | 4 | 6.1 | L | 19 | 2.6 |
| beta (relaxed alert) | 15 | R | 6 | 3.4 | R | 39 | 3.4 | R | 4 | 6.2 | R | 39 | 3 |
| beta (relaxed alert) | 16 | R | 6 | 3.2 | R | 39 | 3.3 | R | 40 | 5 | R | 39 | 2.9 |
| beta (relaxed alert) | 17 | R | 8 | 3.1 | R | 39 | 3.5 | R | 40 | 5.4 | R | 39 | 2.8 |
| beta (relaxed alert) | 18 | R | 8 | 2.8 | R | 7 | 3.2 | R | 40 | 5.4 | R | 39 | 2.6 |
| high beta (aroused and actively engaged) | 19 | R | 8 | 2.7 | R | 7 | 3.3 | R | 40 | 5.6 | R | 19 | 2.7 |
| high beta (aroused and actively engaged) | 20 | R | 40 | 2.4 | R | 7 | 3.7 | R | 40 | 6 | R | 7 | 3.4 |
| high beta (aroused and actively engaged) | 21 | R | 40 | 3.2 | R | 39 | 3.9 | R | 40 | 6.5 | R | 39 | 2.9 |
| high beta (aroused and actively engaged) | 22 | R | 40 | 2.4 | R | 7 | 3.5 | R | 40 | 6.1 | R | 40 | 3.3 |
| high beta (aroused and actively engaged) | 23 | R | 40 | 3.1 | R | 7 | 3.7 | R | 40 | 5.8 | R | 39 | 3.4 |
| high beta (aroused and actively engaged) | 24 | R | 40 | 3 | R | 39 | 3.6 | R | 40 | 5.7 | R | 39 | 3.2 |
| high beta (aroused and actively engaged) | 25 | R | 40 | 2.9 | R | 39 | 3.5 | R | 40 | 4.9 | R | 39 | 3 |
| high beta (aroused and actively engaged) | 26 | R | 40 | 3.3 | R | 39 | 3.5 | R | 40 | 4.4 | R | 39 | 3 |
| high beta (aroused and actively engaged) | 27 | R | 40 | 3 | R | 39 | 3.3 | R | 40 | 4.7 | R | 39 | 2.8 |
| high beta (aroused and actively engaged) | 28 | R | 40 | 3.2 | R | 7 | 3.4 | R | 40 | 5.3 | R | 39 | 2.3 |
| high beta (aroused and actively engaged) | 29 | R | 40 | 3.3 | R | 39 | 3.2 | R | 40 | 4.5 | R | 39 | 2.8 |
| high beta (aroused and actively engaged) | 30 | R | 40 | 3.2 | R | 7 | 3.1 | R | 40 | 4.6 | R | 39 | 2.8 |

**Supplemental Table 7. Z-scores by Brodmann Area by arousal state for Subject 6 (S6)**

| S6 | | Eyes Closed | | | | | | Eyes Open | | | | | |
|---|---|---|---|---|---|---|---|---|---|---|---|---|---|
| | | PRE | | | POST | | | PRE | | | Post | | |
| Arousal State | Freq. (Hz) | Side | Brodmann Area | Z-score | Side | Brodmann Area | Z-score | Side | Brodmann Area | Z-score | Side | Brodmann Area | Z-score |
| Delta (deep sleep) | 1 | L | 6 | 3.2 | R | 13 | 3.2 | R | 8 | 2.3 | L | 13 | 2.2 |
| Delta (deep sleep) | 2 | L | 24 | 3.3 | R | 13 | 3.4 | L | 18 | 2.8 | L | 13 | 2.8 |
| Delta (deep sleep) | 3 | R | 32 | 4 | R | 13 | 3.5 | L | 24 | 3 | L | 11 | 3.1 |
| Delta (deep sleep) | 4 | R | 9 | 4 | R | 40 | 3.7 | L | 9 | 3.5 | L | 11 | 3.7 |
| theta(drowsy) | 5 | R | 9 | 4.2 | R | 1 | 3.5 | R | 9 | 3.6 | L | 11 | 3.9 |
| theta(drowsy) | 6 | L | 21 | 4.1 | R | 2 | 3.4 | R | 46 | 3.8 | L | 11 | 3.7 |
| theta(drowsy) | 7 | L | 47 | 4.4 | L | 47 | 3.2 | L | 38 | 4 | L | 38 | 3.7 |
| alpha(wakeful) | 8 | R | 8 | 3.7 | R | 9 | 2.6 | R | 8 | 3.3 | L | 47 | 2.5 |
| alpha(wakeful) | 9 | R | 9 | 4 | R | 9 | 2.5 | R | 8 | 3.2 | R | 9 | 2.2 |
| alpha(wakeful) | 10 | R | 8 | 4 | R | 8 | 2.3 | L | 8 | 3.4 | R | 9 | 2.1 |
| alpha(wakeful) | 11 | R | 6 | 4.1 | R | 9 | 2.4 | L | 9 | 3.6 | L | 47 | 2.3 |
| beta (relaxed alert) | 12 | R | 8 | 4.1 | R | 8 | 2.6 | L | 9 | 3.8 | R | 9 | 2.5 |
| beta (relaxed alert) | 13 | R | 8 | 3.8 | R | 8 | 2.5 | L | 9 | 3.6 | R | 9 | 2 |
| beta (relaxed alert) | 14 | R | 8 | 3.3 | R | 8 | 2 | R | 9 | 2.8 | L | 3 | 1.8 |
| beta (relaxed alert) | 15 | R | 9 | 3.2 | R | 8 | 1.9 | R | 9 | 2.4 | L | 40 | 1.6 |
| beta (relaxed alert) | 16 | R | 9 | 2.7 | R | 8 | 1.7 | R | 9 | 2 | L | 40 | 1.6 |
| beta (relaxed alert) | 17 | R | 9 | 2.6 | R | 8 | 1.4 | R | 9 | 1.6 | L | 40 | 1.6 |
| beta (relaxed alert) | 18 | R | 9 | 2.4 | R | 8 | 1.3 | R | 9 | 1.4 | L | 40 | 1.6 |
| high beta (aroused and actively engaged) | 19 | R | 8 | 2.1 | L | 40 | 1.3 | R | 9 | 1.2 | L | 40 | 1.5 |
| high beta (aroused and actively engaged) | 20 | R | 9 | 2.2 | R | 40 | 1.2 | R | 9 | 1.3 | L | 40 | 1.2 |
| high beta (aroused and actively engaged) | 21 | R | 8 | 1.9 | R | 40 | 1.2 | L | 9 | 1.3 | L | 39 | 1.2 |
| high beta (aroused and actively engaged) | 22 | R | 8 | 1.8 | L | 7 | 1.3 | L | 20 | 1.3 | L | 19 | 0.8 |
| high beta (aroused and actively engaged) | 23 | R | 8 | 1.8 | L | 7 | 1.3 | R | 38 | 1.5 | L | 19 | 0.9 |
| high beta (aroused and actively engaged) | 24 | R | 8 | 1.8 | R | 40 | 1.3 | R | 38 | 1.4 | L | 7 | 0.9 |
| high beta (aroused and actively engaged) | 25 | R | 8 | 1.7 | L | 7 | 1.5 | R | 38 | 1.3 | L | 19 | 0.7 |
| high beta (aroused and actively engaged) | 26 | L | 7 | 1.8 | L | 7 | 1.7 | L | 7 | 1.1 | R | 40 | 0.9 |
| high beta (aroused and actively engaged) | 27 | L | 7 | 1.9 | L | 7 | 1.7 | L | 7 | 1.2 | L | 7 | 0.8 |
| high beta (aroused and actively engaged) | 28 | L | 7 | 2.1 | L | 7 | 1.7 | L | 7 | 1.1 | L | 7 | 0.9 |
| high beta (aroused and actively engaged) | 29 | R | 8 | 1.8 | L | 7 | 1.7 | L | 7 | 1.2 | L | 7 | 1 |
| high beta (aroused and actively engaged) | 30 | L | 7 | 1.8 | L | 7 | 1.5 | L | 7 | 1.5 | L | 7 | 1 |

**Supplemental Table 8. Z-scores by Brodmann Area by arousal state for Subject 7 (S7)**

| S7 | | Eyes Closed | | | | | | Eyes Open | | | | | |
|---|---|---|---|---|---|---|---|---|---|---|---|---|---|
| | | PRE | | | POST | | | PRE | | | Post | | |
| Arousal State | Freq. (Hz) | Side | Brodmann Area | Z-score | Side | Brodmann Area | Z-score | Side | Brodmann Area | Z-score | Side | Brodmann Area | Z-score |
| Delta (deep sleep) | 1 | L | 24 | 2.5 | L | 3 | 3 | L | 20 | -4.5 | R | 9 | 1.7 |
| Delta (deep sleep) | 2 | L | 23 | 3 | L | 13 | 3.3 | L | 20 | -3.2 | L | 11 | 2.1 |
| Delta (deep sleep) | 3 | L | 24 | 3.1 | L | 3 | 3.8 | L | 13 | -2.8 | L | 11 | 2.5 |
| Delta (deep sleep) | 4 | L | 6 | 3.3 | L | 3 | 4.1 | L | 6 | -3.4 | L | 11 | 2.8 |
| theta(drowsy) | 5 | L | 6 | 2.9 | L | 1 | 4.8 | L | 11 | -3.5 | R | 11 | 2.9 |
| theta(drowsy) | 6 | L | 6 | 2.6 | L | 3 | 4.7 | R | 47 | -3.4 | R | 11 | 2.8 |
| theta(drowsy) | 7 | L | 6 | 2 | L | 1 | 4 | R | 11 | -3.4 | R | 11 | 2.5 |
| alpha(wakeful) | 8 | L | 6 | 1.4 | L | 1 | 3.3 | R | 11 | -2.8 | R | 11 | 1.4 |
| alpha(wakeful) | 9 | R | 6 | 1.2 | L | 1 | 3.2 | L | 46 | -3.4 | R | 11 | 1.5 |
| alpha(wakeful) | 10 | L | 8 | 1.3 | L | 1 | 3.1 | R | 47 | -3.4 | R | 11 | 1.6 |
| alpha(wakeful) | 11 | R | 6 | 1.4 | L | 1 | 3.2 | L | 10 | -3.5 | R | 6 | 1.5 |
| beta (relaxed alert) | 12 | R | 6 | 1.5 | L | 1 | 3.2 | L | 4 | -4.4 | L | 9 | 1.5 |
| beta (relaxed alert) | 13 | R | 6 | 1.6 | L | 1 | 3.6 | L | 44 | -4.6 | R | 6 | 1.6 |
| beta (relaxed alert) | 14 | R | 6 | 1.6 | L | 1 | 3.4 | L | 13 | -3.8 | R | 6 | 1.1 |
| beta (relaxed alert) | 15 | L | 6 | 1.2 | L | 1 | 3.3 | L | 20 | -4.2 | L | 13 | -1 |
| beta (relaxed alert) | 16 | L | 6 | 1 | L | 1 | 2.9 | L | 20 | -3.9 | L | 22 | -1.1 |
| beta (relaxed alert) | 17 | L | 13 | -1 | L | 40 | 2.9 | L | 13 | -3.3 | L | 21 | -1 |
| beta (relaxed alert) | 18 | L | 13 | -1.2 | L | 40 | 2.9 | L | 13 | -3.5 | L | 22 | -1.2 |
| high beta (aroused and actively engaged) | 19 | L | 13 | -1.2 | L | 40 | 3.1 | L | 13 | -3.3 | L | 22 | -1 |
| high beta (aroused and actively engaged) | 20 | L | 13 | -1.3 | L | 40 | 2.9 | L | 13 | -3.2 | R | 13 | -1.1 |
| high beta (aroused and actively engaged) | 21 | L | 13 | -1.1 | L | 40 | 2.8 | L | 13 | -3.2 | L | 13 | -1.1 |
| high beta (aroused and actively engaged) | 22 | L | 13 | -1.1 | L | 40 | 3 | L | 24 | -2.9 | L | 13 | -1.1 |
| high beta (aroused and actively engaged) | 23 | L | 32 | -1.1 | L | 40 | 3.3 | L | 13 | -2.8 | L | 13 | -1.2 |
| high beta (aroused and actively engaged) | 24 | L | 13 | -1.3 | L | 40 | 3.5 | L | 40 | -2.3 | L | 13 | -1.2 |
| high beta (aroused and actively engaged) | 25 | L | 21 | -1.3 | L | 40 | 3.7 | L | 13 | -2.5 | L | 13 | -1.2 |
| high beta (aroused and actively engaged) | 26 | L | 21 | -1.3 | L | 40 | 3.6 | R | 4 | -2.6 | L | 13 | -1.2 |
| high beta (aroused and actively engaged) | 27 | L | 24 | -1.2 | L | 40 | 3.8 | R | 13 | -2.2 | L | 22 | -1.1 |
| high beta (aroused and actively engaged) | 28 | L | 21 | -1.3 | L | 40 | 3.9 | R | 4 | -2.4 | L | 13 | -1.1 |
| high beta (aroused and actively engaged) | 29 | L | 32 | -1.3 | L | 40 | 3.8 | R | 13 | -2.2 | L | 7 | 1.2 |
| high beta (aroused and actively engaged) | 30 | L | 24 | -1.3 | L | 40 | 4 | L | 24 | -2.7 | L | 22 | -1.1 |

### Supplemental Table 9. Z-scores by Brodmann Area by arousal state for Subject 8 (S8)

| S8 | | Eyes Closed | | | | | | Eyes Open | | | | | |
|---|---|---|---|---|---|---|---|---|---|---|---|---|---|
| | | PRE | | | POST | | | PRE | | | Post | | |
| Arousal State | Freq. (Hz) | Side | Brodmann Area | Z-score | Side | Brodmann Area | Z-score | Side | Brodmann Area | Z-score | Side | Brodmann Area | Z-score |
| Delta (deep sleep) | 1 | L | 13 | 4.5 | L | 13 | 4.3 | L | 13 | 4.1 | L | 19 | 3.8 |
| Delta (deep sleep) | 2 | L | 13 | 4.3 | L | 13 | 3.9 | L | 13 | 3.9 | L | 18 | 3.7 |
| Delta (deep sleep) | 3 | L | 6 | 3.9 | R | 19 | 3.6 | L | 13 | 3.9 | L | 19 | 3.7 |
| Delta (deep sleep) | 4 | L | 3 | 4.2 | L | 22 | 4.1 | L | 40 | 3.6 | R | 17 | 3.7 |
| theta(drowsy) | 5 | L | 1 | 4.1 | L | 40 | 4.3 | L | 40 | 3.5 | R | 9 | 3.6 |
| theta(drowsy) | 6 | L | 3 | 4.2 | L | 6 | 4.1 | R | 40 | 3.4 | R | 9 | 3.2 |
| theta(drowsy) | 7 | L | 1 | 3.4 | R | 6 | 2.9 | L | 4 | 2.8 | R | 9 | 3.8 |
| alpha(wakeful) | 8 | L | 1 | 2.5 | R | 9 | 2.7 | L | 3 | 2.3 | R | 9 | 3 |
| alpha(wakeful) | 9 | L | 3 | 2.3 | L | 8 | 3.2 | L | 6 | 2 | R | 8 | 2.4 |
| alpha(wakeful) | 10 | L | 3 | 1.7 | R | 8 | 2.6 | L | 6 | 1.7 | L | 8 | 3.4 |
| alpha(wakeful) | 11 | L | 6 | 1.9 | L | 8 | 2.6 | R | 6 | 1.8 | L | 8 | 3 |
| beta (relaxed alert) | 12 | L | 1 | 2.1 | R | 8 | 2.8 | | | | R | 9 | 2.8 |
| beta (relaxed alert) | 13 | R | 6 | 2.6 | L | 6 | 3 | L | 6 | 2.9 | L | 19 | 2.9 |
| beta (relaxed alert) | 14 | R | 6 | 2.5 | L | 6 | 2.6 | L | 6 | 2.7 | L | 18 | 2.7 |
| beta (relaxed alert) | 15 | L | 40 | 1.8 | R | 6 | 2.5 | L | 6 | 2.5 | L | 18 | 3 |
| beta (relaxed alert) | 16 | L | 6 | 2 | L | 6 | 2.1 | L | 6 | 2.4 | R | 18 | 3.1 |
| beta (relaxed alert) | 17 | L | 40 | 1.7 | L | 20 | 2 | R | 6 | 2.1 | L | 19 | 2.9 |
| beta (relaxed alert) | 18 | R | 6 | 1.7 | L | 21 | 2.2 | L | 6 | 2 | L | 19 | 2.6 |
| high beta (aroused and actively engaged) | 19 | L | 6 | 1.7 | L | 7 | 2 | L | 6 | 2 | L | 19 | 2.2 |
| high beta (aroused and actively engaged) | 20 | L | 40 | 1.5 | L | 7 | 2.2 | L | 6 | 2.9 | L | 19 | 2.4 |
| high beta (aroused and actively engaged) | 21 | L | 6 | 1.9 | R | 40 | 2.3 | L | 6 | 2.9 | L | 18 | 2.5 |
| high beta (aroused and actively engaged) | 22 | L | 6 | 1.9 | R | 40 | 2.2 | L | 5 | 3 | L | 18 | 2.8 |
| high beta (aroused and actively engaged) | 23 | L | 40 | 1.9 | L | 7 | 2.6 | R | 39 | 3 | L | 18 | 3.1 |
| high beta (aroused and actively engaged) | 24 | L | 40 | 2.2 | L | 7 | 3 | R | 39 | 2.9 | L | 18 | 2.7 |
| high beta (aroused and actively engaged) | 25 | L | 40 | 2.2 | R | 40 | 2.7 | R | 39 | 2.6 | L | 18 | 2.7 |
| high beta (aroused and actively engaged) | 26 | L | 40 | 2.1 | L | 7 | 3 | R | 39 | 2.5 | L | 7 | 2.5 |
| high beta (aroused and actively engaged) | 27 | L | 7 | 2 | L | 7 | 3.1 | R | 39 | 2.1 | L | 18 | 2.5 |
| high beta (aroused and actively engaged) | 28 | L | 40 | 2.2 | L | 7 | 3.6 | R | 39 | 2.5 | R | 18 | 2.3 |
| high beta (aroused and actively engaged) | 29 | L | 40 | 2.2 | L | 7 | 3 | R | 19 | 2.3 | R | 18 | 2.6 |
| high beta (aroused and actively engaged) | 30 | L | 40 | 2.3 | L | 7 | 3.2 | L | 5 | 1.9 | R | 18 | 2.3 |

**Supplementary Table 10. Clinically significant z-scores by Brodmann Areas by subject**

| | | EYES CLOSED | | | | | | | | | | EYES OPEN | | | | | | | |
|---|---|---|---|---|---|---|---|---|---|---|---|---|---|---|---|---|---|---|---|
| | | BA 6 | | BA 7 | | BA 8 | | BA 9 | | BA 40 | | BA 4 | | BA 9 | | BA 19 | | BA 39 | |
| Subj. | Channel | Pre | Post | Pre | Post | Pre | Post | Pre | Post | Pre | Post | Pre | Post | Pre | Post | Pre | Post | Pre | Post |
| S1 | 1 (delta) | 2.3 | - | | | | | | | | | | | | | | | | |
| | 7 (theta) | | | | | | | | | | | 2.7 | - | | | | | | |
| | 11 (alpha) | | | | | | | | | | | | | 3.1 | - | | | | |
| | 15 (beta) | | | | | | | 2.3 | - | | | | | | | | | | |
| | 23 (high beta) | | | | | 1.6 | - | | | | | | | | | | | | |
| S2 | 5 (theta) | | | | | | | | | - | 4.0 | | | | | | | | |
| | 11 (alpha) | | | | | | | | | | | | | 2.4 | - | | | | |
| | 15 (beta) | | | | | | | 2.3 | - | | | | | | | | | | |
| | 19 (high beta) | | | | | 1.5 | - | | | - | 1.6 | | | | | | | | |
| | 21 (high beta) | | | | | | | | | - | 1.8 | | | | | | | - | 1.8 |
| | 23 (high beta) | | | | | 1.5 | - | | | | | | | | | | | | |
| | 24 (high beta) | | | | | 1.5 | - | | | | | | | | | | | | |
| | 30 (high beta) | | | - | 1.5 | | | | | | | | | | | | | | |

**Supplementary Table 10. Clinically significant z-scores by Brodmann Areas by subject**

| | | EYES CLOSED | | | | | | | | | | EYES OPEN | | | | | | | |
|---|---|---|---|---|---|---|---|---|---|---|---|---|---|---|---|---|---|---|---|
| | | BA 6 | | BA 7 | | BA 8 | | BA 9 | | BA 40 | | BA 4 | | BA 9 | | BA 19 | | BA 39 | |
| Subj. | Channel | Pre | Post | Pre | Post | Pre | Post | Pre | Post | Pre | Post | Pre | Post | Pre | Post | Pre | Post | Pre | Post |
| S4 | 7 (theta) | | | | | | | | | | | 3.0 | - | | | | | | |
| | 19 (high beta) | | | | | | | | | - | 2.2 | | | | | | | | |
| | 21 (high beta) | | | | | | | | | - | 2.4 | | | | | | | | |
| | 29 (high beta) | | | 2.2 | 2.7 | | | | | | | | | | | | | | |
| | 30 (high beta) | | | 2.5 | 2.8 | | | | | | | | | | | | | | |
| S5 | 5 (theta) | | | | | | | | | - | 4.3 | | | | | | | | |
| | 19 (high beta) | | | | | 2.7 | - | | | | | | | | | - | 2.7 | | |
| | 21 (high beta) | | | | | | | | | 3.2 | - | | | | | | | - | 2.9 |
| | 22 (high beta) | | | - | 3.5 | | | | | | | | | | | | | | |
| | 30 (high beta) | | | - | 3.1 | | | | | | | | | | | | | | |
| S6 | 1 (delta) | 3.2 | - | | | | | | | | | | | | | | | | |
| | 11 (alpha) | | | | | | | | | | | | | 3.6 | - | | | | |

**Supplementary Table 10. Clinically significant z-scores by Brodmann Areas by subject**

| | | EYES CLOSED | | | | | | | | | | EYES OPEN | | | | | | | |
|---|---|---|---|---|---|---|---|---|---|---|---|---|---|---|---|---|---|---|---|
| | | BA 6 | | BA 7 | | BA 8 | | BA 9 | | BA 40 | | BA 4 | | BA 9 | | BA 19 | | BA 39 | |
| Subj. | Channel | Pre | Post | Pre | Post | Pre | Post | Pre | Post | Pre | Post | Pre | Post | Pre | Post | Pre | Post | Pre | Post |
| S6 | 15 (beta) | | | | | | | 3.2 | - | | | | | | | | | | |
| | 19 (high beta) | | | | | 2.1 | - | | | | | | | | | | | | |
| | 23 (high beta) | | | | | 1.8 | - | | | | | | | | | | | | |
| | 24 (high beta) | | | | | 1.8 | - | | | | | | | | | | | | |
| | 29 (high beta) | | | - | 1.7 | | | | | | | | | | | | | | |
| | 30 (high beta) | | | 1.8 | 1.5 | | | | | | | | | | | | | | |
| S7 | 19 (high beta) | | | | | | | | | - | 3.1 | | | | | | | | |
| | 21 (high beta) | | | | | | | | | - | 2.8 | | | | | | | | |
| S8 | 5 (theta) | | | | | | | | | - | 4.3 | | | | | | | | |
| | 7 (theta) | | | | | | | | | | | 2.8 | - | | | | | | |
| | 19 (high beta) | | | | | | | | | | | | | | | - | 2.2 | | |
| | 21 (high beta) | | | | | | | | | - | 2.3 | | | | | | | | |
